# Auditory tests for characterizing hearing deficits in listeners with various hearing abilities: The BEAR test battery

**DOI:** 10.1101/2020.02.17.20021949

**Authors:** Raul Sanchez-Lopez, Silje Grini Nielsen, Mouhamad El-Haj-Ali, Federica Bianchi, Michal Fereczkowski, Oscar M Cañete, Mengfan Wu, Tobias Neher, Torsten Dau, Sébastien Santurette

## Abstract

The Better hEAring Rehabilitation (BEAR) project aims to provide a new clinical profiling tool – a test battery – for hearing loss characterization. Whereas the loss of sensitivity can be efficiently measured using pure-tone audiometry, the assessment of supra-threshold hearing deficits remains a challenge. In contrast to the classical ‘attenuation-distortion’ model, the proposed BEAR approach is based on the hypothesis that the hearing abilities of a given listener can be characterized along two dimensions, reflecting independent types of perceptual deficits (distortions). A data-driven approach provided evidence for the existence of different auditory profiles with different degrees of distortions. Ten tests were included in a test battery, based on their clinical feasibility, time efficiency and related evidence from the literature. The tests were divided into six categories: audibility, speech perception, binaural processing abilities, loudness perception, spectro-temporal modulation sensitivity and spectro-temporal resolution. Seventy-five listeners with symmetric, mild-to-severe sensorineural hearing loss were selected from a clinical population. The analysis of the results showed interrelations among outcomes related to high-frequency processing and outcome measures related to low-frequency processing abilities. The results showed the ability of the tests to reveal differences among individuals and their potential use in clinical settings.

## 1 INTRODUCTION

In current clinical practice, hearing loss is diagnosed mainly on the basis of pure-tone audiometry (ISO 8253-1, 2010). The audiogram helps differentiate between conductive and sensorineural hearing losses and can characterize the severity of the hearing loss from mild to profound. However, the pure-tone audiogram only assesses the sensitivity to simple sounds, which is not necessarily related to listening abilities at supra-threshold sound pressure levels (e.g. a person’s ability to discriminate speech in noise)

Pure-tone audiometry is often complemented by speech audiometry (ISO 8253-3, 2012), which is a test typically performed in the form of word recognition performance in quiet (Anderson et al., 2018). Although this test can provide information about supra-threshold deficits (Gelfand, 2009), measurements of speech understanding in noise have been found more informative (Killion et al., 2004; Nilsson et al., 1994). Since improving speech intelligibility is usually the main goal of successful hearing rehabilitation, several auditory factors affecting speech intelligibility in noise have been investigated (e.g. Glasberg and Moore, 1989; Houtgast and Festen, 2008; Strelcyk and Dau, 2009). Audibility (in conditions with fluctuating maskers), frequency selectivity (in conditions with stationary noise), and temporal processing acuity (in conditions with speech interferers), have been identified as important factors affecting speech reception thresholds in noise when using meaningful sentences as speech material (e.g. Desloge et al., 2017; Johannesen et al., 2016; Oxenham and Simonson, 2009; Rhebergen et al., 2006)^2^ Thus, a hearing evaluation that goes beyond pure-tone sensitivity and speech intelligibility in quiet would be expected to provide a more accurate characterization of a listener’s hearing deficits.

In Denmark, the Better hEAring Rehabilitation (BEAR) project was initiated with the aim of developing new diagnostic tests and hearing-aid compensation strategies for audiological practice. Although the assessment of individual hearing deficits can be complex, new evidence suggests that the perceptual consequences of a hearing loss can be characterized effectively by two types of hearing deficits, defined as “auditory distortions” (Sanchez-Lopez et al., 2018). By analysing the outcomes of two previous studies (Johannesen et al., 2016; Thorup et al., 2016) with a data-driven approach, Sanchez-Lopez et al. (2018) identified high-frequency hearing loss as the main predictor of one of the distortions, whereas the definition of the second type of distortion was inconclusive. The inconclusiveness in the prediction of the second distortion was most likely due to differences between the two studies in terms of hearing loss profiles and outcome measures. Here, a new dataset was therefore collected based on a heterogeneous group of listeners with audiometric hearing losses ranging from very mild to severe and with a large range of audiometric profiles. To that end, the most informative tests resulting from the analysis of Sanchez-Lopez et al. (2018) were included, together with additional auditory tests that had shown potential for hearing profiling in other previous studies. The tests included in the current study are referred to as the BEAR test battery.

The characterization of hearing deficits beyond the audiogram was considered in several earlier studies (e.g., Lecluyse et al., 2013; Brungart et al., 2014; Rönnberg et al., 2016; Santurette and Dau, 2012; Saunders et al., 1992; Vlaming et al., 2011). Among them, the HEARCOM project (Vlaming et al., 2011) proposed an extended hearing profile formed by the results of several behavioural tests. These tests targeted various auditory domains, such as audibility, loudness perception, speech perception, binaural processing, and spectro-temporal resolution, as well as a test of cognitive abilities. Importantly, while the auditory domains considered in the BEAR test battery are similar to the ones considered in the HEARCOM project, the BEAR project aims to additionally classify the patients in subcategories and to create a link between hearing capacities and hearing-aid parameter settings.

The tests included in the BEAR test battery were chosen based on the following criteria: 1) There is evidence from the hearing research literature that the considered test is informative (i.e., it provides information about the individual hearing deficits) and reliable (i.e., the result of the test does not vary over time); 2) The outcomes of the test may be linked to a hearing-aid fitting strategy; 3) The outcome measures are easy to interpret and to explain to the patient; 4) The task is reasonably time-efficient or can be suitably modified to meet this requirement (e.g., by changing the test paradigm or developing an out-of-clinic solution); 5) The test implementation can be done with equipment available in clinics; 6) The tasks are not too demanding for patients and clinicians; 7) Tests with several outcome measures are prioritized, and 8) The language independent tests are also prioritized.

The selected test battery included measures of audibility, loudness perception, speech perception, binaural processing abilities, spectro-temporal modulation (STM) sensitivity and spectro-temporal resolution. It was implemented and tested in older listeners with different hearing abilities (from mild to severe hearing losses). The goals of the study were: 1) To collect reference data from a representative sample of HI listeners for each of the selected tests, 2) to analyse the test-retest reliability of these tests, 3) to analyse the relationships between the different outcome measures, and 4) to propose a version of the test battery that can be implemented in hearing clinics.

## 2 OVERVIEW OF THE TEST BATTERY

The test battery consisted of ten tests (9 tests besides the pure-tone audiometry). The outcomes of the proposed tests are divided into six categories. Table 1 shows the tests and the corresponding auditory domains or categories. By convenience, the domains spectro-temporal modulation sensitivity and spectro-temporal resolution are together in the category spectro-temporal processing. The following sections introduce the experimental methods and present all tests individually. The dataset is publicly available in a Zenodo repository (Sanchez-Lopez et al., 2019). More details about the method can be found in the supplementary material in the data repository.

**Table 1.**
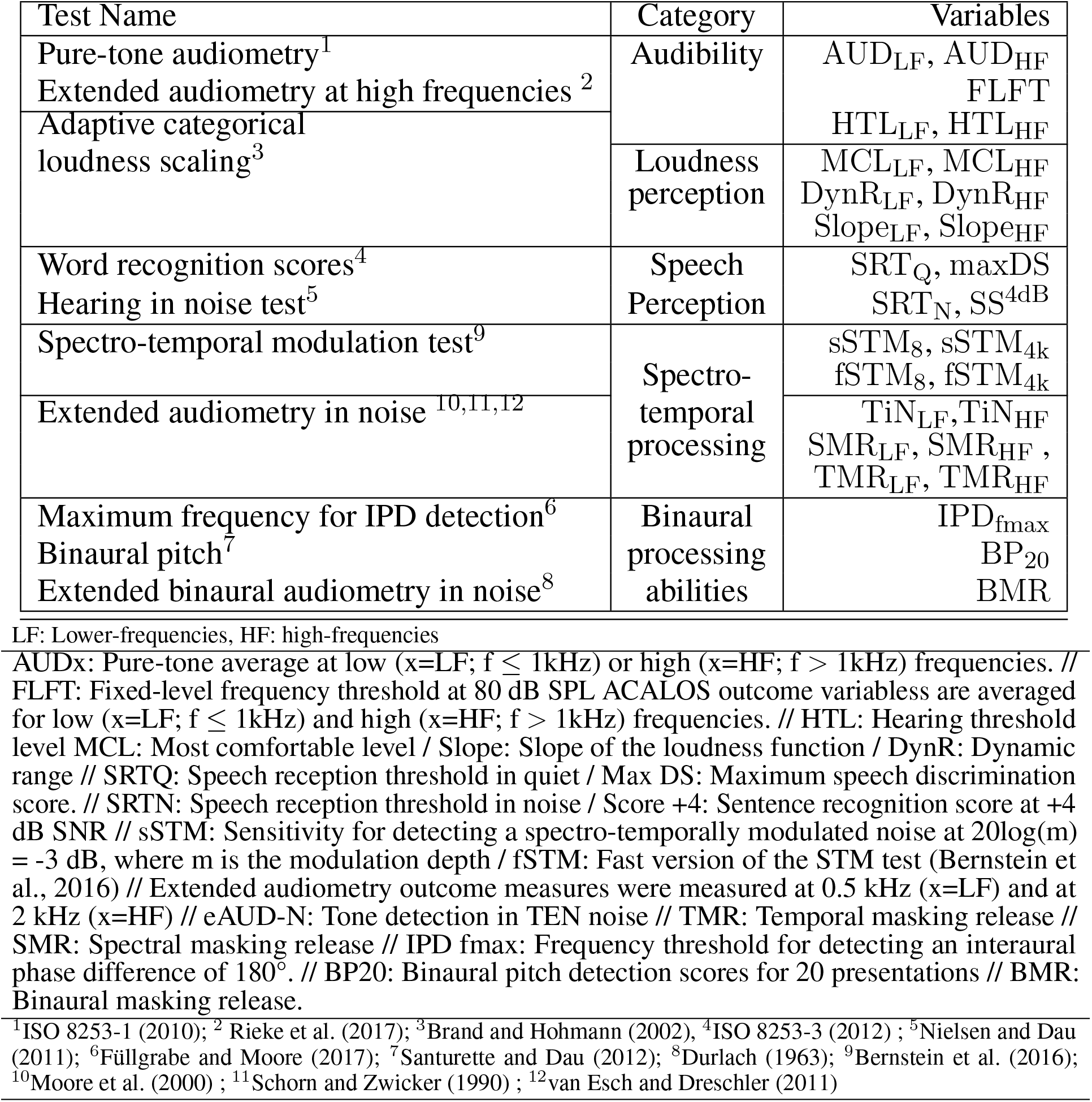
List of the tests included in the BEAR test battery and their corresponding auditory domains.

### 2.1 Reference data from younger normal-hearing listeners

Although many of the tests included in the test battery are based on previous studies with normative data, a group of 11 young normal-hearing listeners were tested in the facilities of the Technical University of Denmark (DTU) and the University of Southern Denmark (SDU) to obtain reference data for this specific implementation of each of the tests. The summary statistics of the outcome variables from Table 1 are shown in Table 2.

**Table 2.** Reference data of the young normal-hearing group. The data is shown as the 5th, 25th, 50th, 75th and 95th percentiles. In tests performed monoaurally, the summary corresponds to the data of both ears merged together (i.e., 22 ears).

| Outcome | 5th | 25th | 50th | 75th | 95th | Unit |
| --- | --- | --- | --- | --- | --- | --- |
| AUD<sub>LF</sub> | -5 | 0 | 0 | 5 | 10 | dB HL |
| AUD<sub>HF</sub> | -10 | 0 | 5 | 10 | 10 | dB HL |
| FLFT | 14.42 | 15.60 | 17.88 | 18.02 | 19.31 | kHz |
| HTL<sub>LF</sub> | -7.5 | -2.5 | 1.25 | 2.5 | 7.5 | dB HL |
| HTL<sub>HF</sub> | -5.5 | 2.5 | 8.75 | 15 | 23.5 | dB HL |
| MCL<sub>LF</sub> | 55 | 62.5 | 70 | 72.5 | 78 | dB HL |
| MCL<sub>HF</sub> | 62.5 | 70 | 75 | 80 | 87.5 | dB HL |
| DynR<sub>LF</sub> | 87.5 | 92.5 | 97.5 | 105 | 117.5 | dB |
| DynR<sub>HF</sub> | 74.5 | 85 | 92.5 | 100 | 105 | dB |
| Slope<sub>LF</sub> | 0.27 | 0.30 | 0.33 | 0.37 | 0.42 | CU/dB |
| Slope<sub>HF</sub> | 0.27 | 0.30 | 0.33 | 0.37 | 0.44 | CU/dB |
| SRT<sub>Q</sub> | 6 | 10 | 14 | 18 | 21 | dB SPL |
| maxDS | 96.7 | 100 | 100 | 100 | 100 | % Corr |
| SRT<sub>N</sub> | -3.48 | -1.84 | -0.85 | -0.19 | 1 | dB SNR |
| SS<sub>+4dB</sub> | 80 | 90 | 95 | 100 | 100 | % Corr |
| sSTM<sub>8</sub> | 2.40 | 3.07 | 3.07 | 3.07 | 3.07 | d' |
| sSTM<sub>4k</sub> | 0.30 | 2.40 | 3.07 | 3.07 | 3.07 | d' |
| fSTM<sub>8</sub> | -13.5 | -12.7 | -12.3 | -10.5 | -6.1 | dB ML |
| fSTM<sub>4k</sub> | -9.1 | -6.5 | -4.375 | -3.375 | -2.7 | dB ML |
| TiN<sub>LF</sub> | 64.25 | 66.5 | 68.25 | 69.5 | 73.6 | dB HL |
| TiN<sub>HF</sub> | 64.1 | 68 | 69.375 | 70 | 73.85 | dB HL |
| TMR<sub>LF</sub> | 3.55 | 7.4167 | 9.375 | 13 | 15.95 | dB |
| TMR<sub>HF</sub> | 3.1 | 8.25 | 10.875 | 13 | 18.15 | dB |
| SMR<sub>LF</sub> | 16.55 | 19.75 | 21.75 | 23.25 | 26.15 | dB |
| SMR<sub>HF</sub> | 19.6 | 26 | 28.5833 | 30.75 | 31.25 | dB |
| IPD | 0.86 | 1.20 | 1.22 | 1.29 | 1.40 | kHz |
| BP<sub>20</sub> | 50 | 98.75 | 100 | 100 | 100 | % Corr |
| BMR | 14.3 | 15.875 | 17 | 18.4375 | 22.1 | dB |

### 2.2 Time efficiency of the test battery

The examiners kept track of the time used by each of the participants in completing the test battery. In the case of unexpected events (e.g., unexpected or incongruent results), these events were cautiously annotated for later investigation. Regarding the test procedure, additional repetitions of the threshold estimations were needed if: 1) a repetition was considered as an outlier if a given threshold was greater than three scaled median absolute deviations of the two repetitions; or 2) the responses of the listeners during the tracking procedure were inconsistent or reached the maximum or minimum possible values. In that case, the measurement was considered an invalid or “missing” data point.

The timing annotated of the individual tests are shown in Figure 1. Besides, the probability of needing an additional measurement and mean number of extra repetitions per listener are shown in Table 3. The repetitions were only suggested when the test was done using the alternaltive-forced choice (AFC) framework (i.e., the IPD test, the STM test and the eAUD test in all the conditions). The total testing time was approximately 2’5 hours excluding the initial interview, information about the study and preparations.

**Table 3.**
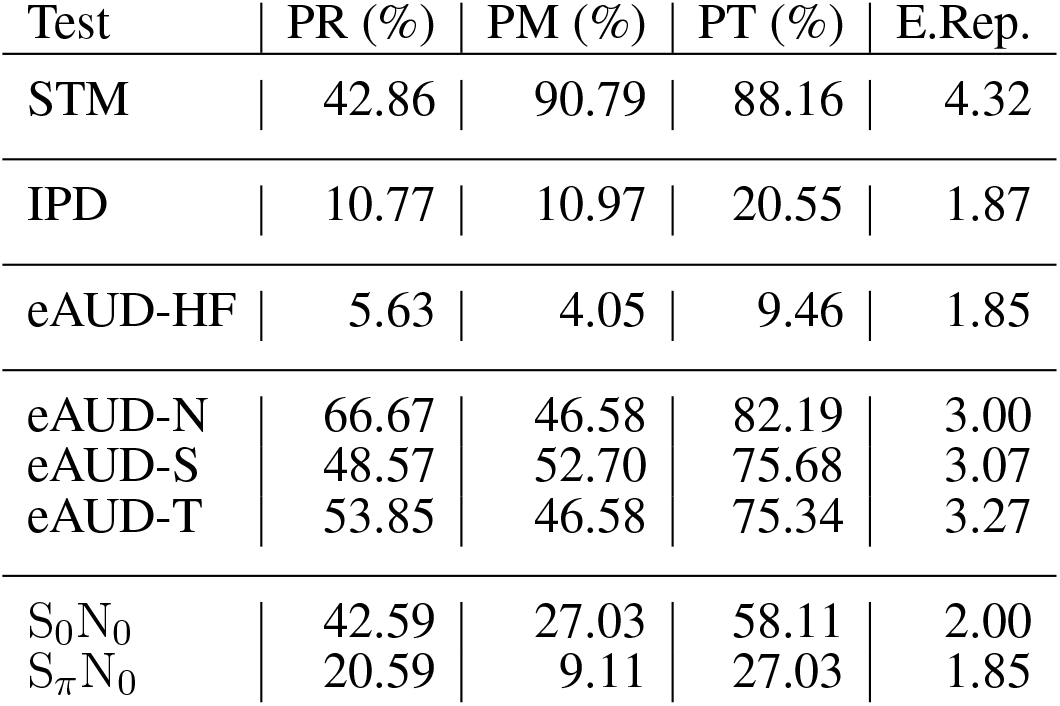
Table with the probability of needing repetitions (PR), and the probability of having missing values (PM). The total probability of repetitions (PT). The mean number of extra repetitions (E. Rep).

| Test | PR (%) | PM (%) | PT (%) | E.Rep. |
| --- | --- | --- | --- | --- |
| STM | 42.86 | 90.79 | 88.16 | 4.32 |
| IPD | 10.77 | 10.97 | 20.55 | 1.87 |
| eAUD-HF | 5.63 | 4.05 | 9.46 | 1.85 |
| eAUD-N | 66.67 | 46.58 | 82.19 | 3.00 |
| eAUD-S | 48.57 | 52.70 | 75.68 | 3.07 |
| eAUD-T | 53.85 | 46.58 | 75.34 | 3.27 |
| S<sub>0</sub>N<sub>0</sub> | 42.59 | 27.03 | 58.11 | 2.00 |
| S<sub>π</sub>N<sub>0</sub> | 20.59 | 9.11 | 27.03 | 1.85 |

**Figure 1.**
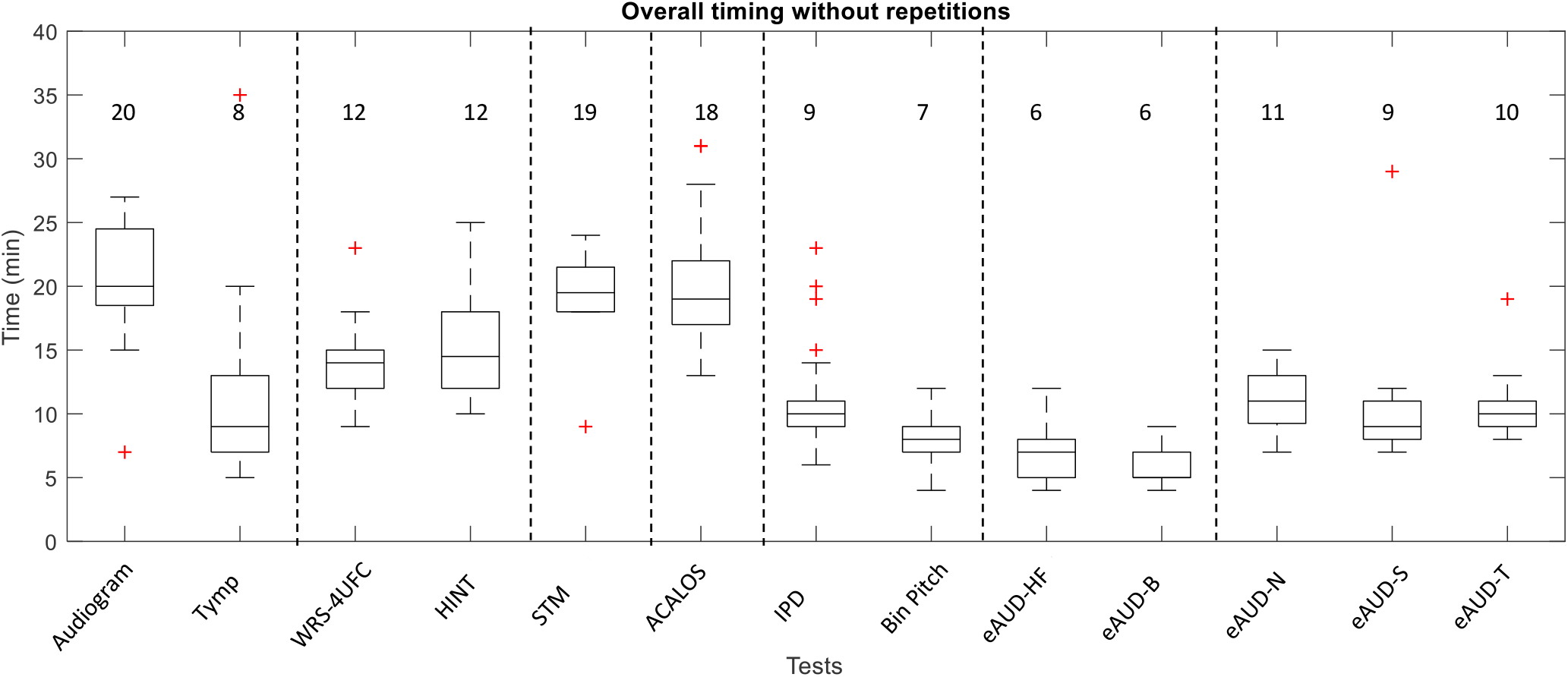
The overall time of the different tests in the test battery including the instructions. The data corresponds to the annotations of the examiners. The basic examination with the audiometry and the tympanometry (Tymp) are included. The numbers represent the rounded median in minutes.

## 3 GENERAL METHODS

### 3.1 Participants and general setup

Seventy-five listeners (38 of them females) participated in the study, who were aged between 59 and 82 years (median: 71 years). Five participants were considered older normal hearing (ONH) with thresholds below 25 dB Hearing Level (HL) in the frequency range between 0.25 and 4 kHz in both ears and no larger than 40 dB HL at 8kHz (PTA *≤* 22 dB HL)^3^. PTA was defined as the pure-tone average between 0.5, 1 and 2 kHz as it is typically reported (Vermiglio et al., 2020). Two of these participants were not usual hearing-aid users. The hearing-impaired listeners (HI) group consisted of 70 participants with symmetric sensorineural hearing losses. Symmetric sensorineural hearing loss was defined as an interaural difference (ID) *≤* 15 dB HL at frequencies below 8 kHz and ID *≤* 25 dB HL at 8 kHz and air-bone gap *≤* 10 dB HL. The pure-tone audiograms of the participants are shown in Figure 2. The participants eligible for the present study had audiometric thresholds *≤* 55 dB HL (pure-tone audiometry not older than 1 year) in the range between 125 and 1000 Hz. Participants with a pure tone threshold *≥* 75 dB HL at 2 kHz were excluded from the study as it was unlikely that it would be feasible to perform all the tests due to audibility issues.

**Figure 2.**
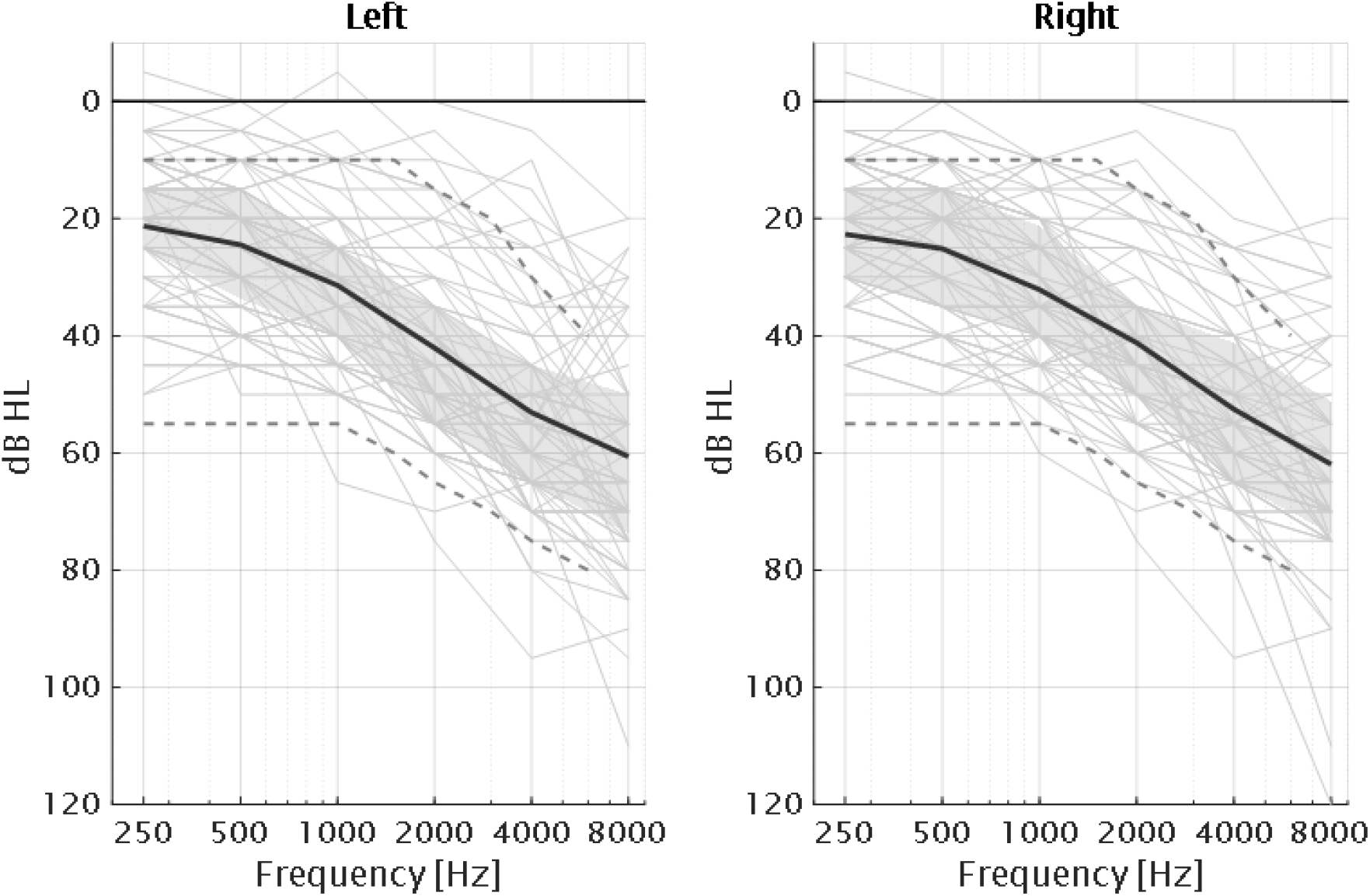
Audiograms of the 75 participants of the study together with the average for each ear (dark solid lines) and interquartile ranges (grey areas). The grey dashed lines correspond to the standard audiograms N1 and N4 from Bisgaard et al. (2010).

The participants were recruited from the BEAR database (Houmoller et al., 2021) at Odense University Hospital (OUH), from the patient database at Bispebjerg Hospital (BBH), and from the database at the Hearing Systems Section at the Technical University of Denmark (DTU). The study was approved by the Science-Ethics Committee for the Capital Region of Denmark, H-16036391. All participants gave written informed consent and some of them received economical compensation for their participation depending on the test site and whether the participant was willing to particpate without compensation.

### 3.2 Equipment

The basic audiological assessment consisted of pure-tone audiometry, wideband tympanometry (Rosowski et al., 2013) and middle ear muscle reflex, and was conducted in the facilities of OUH, BBH and DTU. The rest of the tests were performed via PC in a double-walled sound-insulated booth (BBH and DTU) or in a small anechoic chamber (OUH). The tests were implemented in Matlab with a graphical user interface (GUI) that the examiner could operate without programming experience. Most of the tests were implemented using a modular framework for psychoacoustic experiments (AFC; Ewert, 2013), except for HINT, provided by Jens Bo Nielsen and Binaural Pitch test which was a reimplementation of the Binaural Pitch Test v1.0, Bispebjerg hospital, 2008. The participants were seated in the room and the stimuli were presented through headphones (Sennheiser HDA200) connected to a headphone-amplifier (SPL phonic) and an audio interface (RME Surface 24-bit). The equipment was calibrated using an artificial ear according to IEC 60318-1:2009. The tests consisting of threshold estimation using the AFC framework were repeated at least two times and the mean of the two measurements was considered as the final value. To ensure the quality of the data collected, a repetition was considered as an outlier if it was greater than three scaled median absolute deviations and additional repetitions were suggested by the framework until a certain standard deviation across measures was achieved.

### 3.3 Analysis of test reliability

The test-retest reliability of the test battery was assessed using intraclass correlation coefficients (ICC; Koo and Li, 2016), and the standard error of measurement (SEM; Stratford and Goldsmith, 1997). It was of special interest to test the reliability in older listeners with different hearing abilities. Therefore, test-retest measurements were performed with a subgroup consisting of 11 participants for all tests of the test battery. The seven listeners had bilateral hearing loss with a mean PTA of 31 dB HL. The participants were aged between 59 and 82 years (median 69 years). The retest session was conducted within four months after the first visit.

## 4 HIGH-FREQUENCY AUDIBILITY

Recently, elevated thresholds at high frequencies (*>* 8 kHz) have been linked to the concept of “hidden hearing loss” and synaptopathy (Liberman et al., 2016). However, the measurement of audiometric thresholds above 8 kHz is not part of the current clinical practice. The fixed-level frequency threshold (FLFT) has been proposed as a quick and efficient alternative to high-frequency audiometry (Rieke et al., 2017; Prendergast et al., 2020). The test is based on the detection of a tone presented at a fixed level. The frequency of the tone is varied towards high frequencies and the maximum audible frequency at the given level is estimated in an adaptive procedure. Here, a modified version of FLFT, using warble tones presented at 80 dB SPL, was used as the extended audiometry at high frequencies (eAUD-HF).

### 4.1 Method

The procedure used here was a yes/no task using a SIAM procedure (Kaernbach, 1990). As in traditional up-down procedures, the target can be presented in a given trial or not. If the target was detected, the frequency of the warble tone was increased according to a given step size; if it was not detected, the frequency was decreased. For each run, the first two reversals were discarded, and the threshold of each trial was calculated as the average of the four subsequent reversals. This procedure includes catch trials where no sound was presented.

### 4.2 Results and discussion

The results of the FLFT measured at 80 dB SPL are shown in Table 4.

**Table 4.**
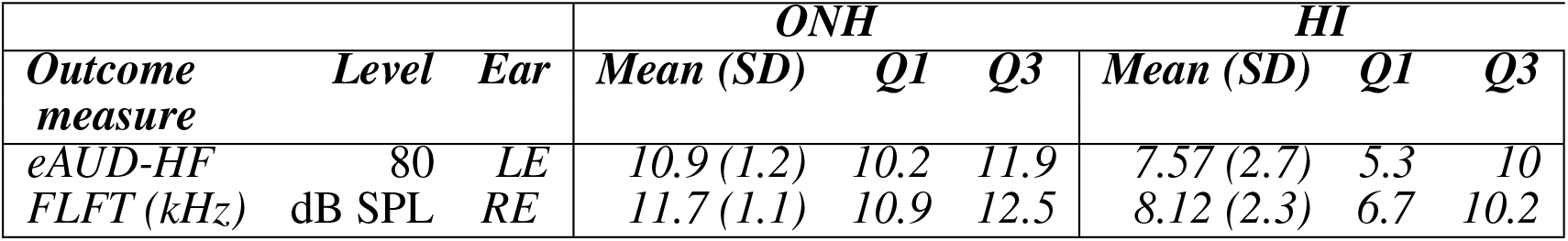
Summary of the results of the Extended Audiometry for high frequencies eAUD-HF. The results are shown as the mean, standard deviation, first and third quantiles (25th and 75th percentiles) for each ear.

The maximum frequency threshold for a tone presented at 80 dB SPL (eAUD-HF) was 11 kHz for the ONH listeners and 8 kHz for the HI listeners. The HI group showed larger variability compared to the NH group (interquartile range: 6 kHz vs. 10 kHz). The eAUD-HF test showed very good reliability (ICC = 0.89; SEM = 495 Hz). These results suggest that the FLFT paradigm might be a good time-efficient alternative to the traditional audiometry for measuring high-frequency sensitivity. A recent study pointed out the importance of off-frequency listening and the role of the excitation of the basal cochlea when presenting narrow-band stimuli in high levels (Encina-Llamas et al., 2019). Knowing the hearing sensitivity at high-frequencies of a given patient might be crucial for better understanding of their supra-threshold deficits. Moreover, The eAUD-HF can include different levels and be a useful not only for ototoxicity monitoring but also in association with other supra-threshold measues.

## 5 LOUDNESS PERCEPTION

Loudness perception can substantially differ between NH and HI listeners and has been connected to the peripheral non-linearity (e.g. Jü rgens et al., 2011). While the growth of loudness shows a non-linear behaviour in a healthy ear, the results from HI listeners suggest that loudness perception becomes linear when outer-hair cell (OHC) function is affected (e.g Moore, 2007). Besides, the possibilities of characterizing hearing deficits, loudness function can be used for fitting hearing aids (e.g. Oetting et al., 2018). Adaptive categorical loudness scaling (ACALOS; Brand and Hohmann, 2002) is the reference method for the current standard (ISO 16832, 2006) for loudness measurements.

### 5.1 Methods

According to the ACALOS method, 1/3-octave band noise were presented sequentially, and the participant had to judge the perceived loudness using a 11-category scale ranging from “not heard” to “extremely loud”. The presentation level of the next stimulus was calculated based on the previous trials. The raw results, which correspond to categorical units (CU) spanned between 0 and 50, were fitted to a model of loudness as described in (Oetting et al., 2014). The outcome measures of the ACALOS presented here are the most comfortable level (MCL), the slope of the loudness function (Slope), and the dynamic range (DynR) defined as the difference between the uncomfortable level (50 CU) and the hearing threshold (0.5 CU). Low-frequency (LF) average corresponds to frequencies below 1.5 kHz, high-frequency (HF) average correspond to frequencies above 1.5 kHz.

### 5.2 Results and discussion

The results of the ACALOS outcome measures are shown in Table 5. The hearing thresholds (HTL) estimated by ACALOS were significantly correlated with the pure-tone audiometric thresholds (*ρ* = 0.88; *p <<* 0.0001) even when looking at the HI group alone (*ρ* = 0.83; *p <<* 0.0001) despite the use of different stimuli and procedure.

**Table 5.** Summary of the results of the adaptive categorial loudness scaling test (ACALOS). The results of the most comfortable level (MCL), slope of the growth of loudness (Slope) and dynamic range (DynR) are shown for low (LF) and high frequencies (HF) as the mean, standard deviation, first and third quantiles (25th and 75th percentiles) for each ear.

| Outcome measure | Freq. Range | Ear | ONH |  |  | HI |  |  |
| --- | --- | --- | --- | --- | --- | --- | --- | --- |
|  |  |  | Mean (SD) | Q1 | Q3 | Mean (SD) | Q1 | Q3 |
| MCL (dB HL) | LF | LE | 81.5 (14.8) | 73.3 | 84.1 | 80.6 (8.4) | 76.4 | 85.8 |
|  |  | RE | 76.5 (13.2) | 70 | 80 | 79.1 (7.9) | 74.7 | 84.1 |
|  | HF | LE | 79.0 (17.6) | 66.6 | 90.8 | 82.7 (12.3) | 75.8 | 90 |
|  |  | RE | 73.8 (17.2) | 65 | 80 | 80.3 (9.9) | 74.7 | 87.5 |
| Slope (CU/dB) | LF | LE | 0.35 (0.1) | 0.3 | 0.4 | 0.45 (0.1) | 0.3 | 0.5 |
|  |  | RE | 0.36 (0.1) | 0.3 | 0.4 | 0.48 (0.2) | 0.3 | 0.5 |
|  | HF | LE | 0.45 (0.1) | 0.3 | 0.4 | 0.84 (0.5) | 0.5 | 0.9 |
|  |  | RE | 0.41 (0.1) | 0.3 | 0.4 | 0.81 (0.4) | 0.5 | 0.9 |
| DynR (dB HL) | LF | LE | 91.5 (16.8) | 78.3 | 97.5 | 76.7 (15.8) | 64.5 | 88.3 |
|  |  | RE | 91.1 (18.8) | 79.1 | 100 | 73.9 (16.0) | 61.6 | 86.8 |
|  | HF | LE | 77.6 (18.2) | 72.5 | 85.8 | 50.8 (15.1) | 40.6 | 60.2 |
|  |  | RE | 78.6 (17.9) | 67.5 | 90.8 | 50.7 (15.5) | 38.9 | 60.4 |

The average MCL estimate ranged between 73 and 82 dB HL in both groups and for both frequency ranges. The average slope of the loudness growth was slightly steeper for the HI listeners in the low-frequency range (0.45 CU/dB for HI vs. 0.35 CU/dB for ONH) and substantially steeper in the high-frequency range (0.8 CU/dB for HI vs 0.45 CU/dB for NH). The average dynamic range was between 80 and 90 dB HL for the ONH listeners, and smaller for the HI listeners, especially at high frequencies (50.8 dB). Regarding the test-retest reliability, ACALOS showed an excellent reliability for estimating the hearing thresholds (ICC = 0.94; SEM = 4.5 dB), good reliability for estimating the MCL (ICC = 0.68, SEM = 6.5 dB) and very good reliability for estimating the slope (ICC = 0.82; SEM = 0.07 CU/dB). Overall, these results supported the inclusion of ACALOS in a clinical test battery, as it provides several outcomes (hearing thresholds, growth of loudness, MCL and dynamic range). ACALOS also showed a high time efficiency (around 10 min. per ear).

## 6 SPEECH PERCEPTION IN QUIET

### 6.1 Method

The word recognition score with four unforced choices (WRS-4UFC) test was proposed as a systematic and self-administered procedure that allows the estimation of supra-threshold deficits in speech perception in quiet. The speech material was the same as the one used for standard speech audiometry (Dantale I; Elberling et al., 1989) in Danish. The self-administered procedure consisted of the presentation of one word were the patient has to answer in a 4-unforced-choice paradigm (4UFC). After the acoustical presentation of each word, the target written word was assigned randomly to one of four buttons shown to the patient. The other three buttons contained words that were also taken from the Dantale-I corpus. They were chosen based on the lowest Levenshtein phonetic distance (Sanders and Chin, 2009) from the target. The term “unforced” corresponds to an additional choice, a question mark, that the listener can press if none of the four options are considered the right answer. Four lists of 25 words were presented at 40, 30, 20 and 10 dB above the individual PTA, in this order. A logistic function was fitted to the results from each individual ear and the speech reception threshold (SRT_Q_), at which 50% of the words were recognized, and maximum speech discrimination score (maxDS), which was the maximum value of the function. Both outcomes were estimated using psignifit 4 software (Schütt et al., 2016).

### 6.2 Results and discussion

The results of the WRS-4UAFC outcome measures are shown in Table 6.

**Table 6.**
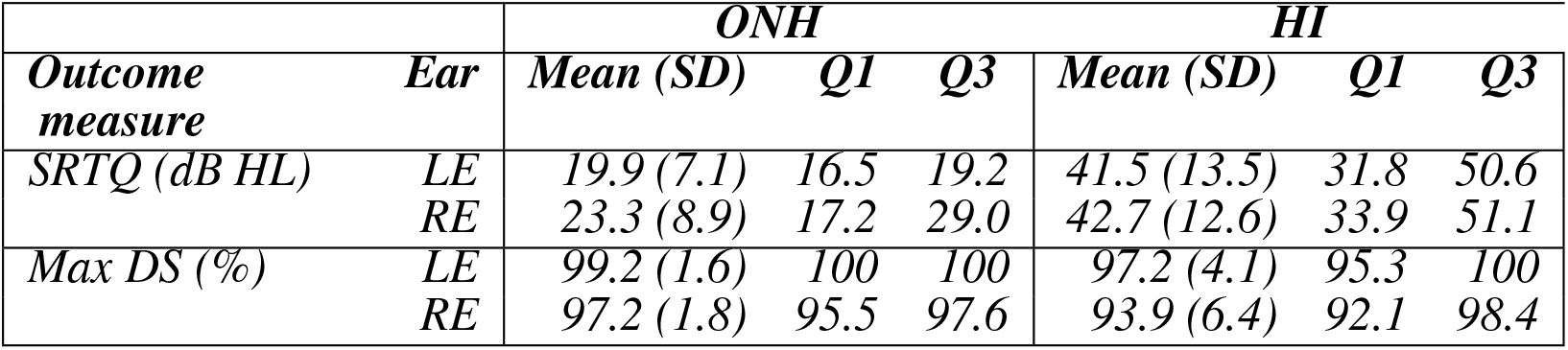
Summary of the results of the word recognition scores (WRS-4UFC) test. The results of the speech recognition threshold (SRT_Q_), and maximum discrimination score (maxDS) are shown as the mean, standard deviation, first and third quantiles (25th and 75th percentiles) for each ear.

The HI listeners’ SRT_Q_ were, on average, 20 dB higher than the ones of the ONH group. The interquartile range for the HI group was about 19 dB whereas for the ONH group it was 3 dB for the left ear (LE) and 11.8 dB for the right ear (RE). The maxDS for both groups was close to 100%. However, the HI listeners showed larger variability, especially in the right ear (SD= 6.42%). In the analysis of the test-retest variability, the WRS-4UFC test showed poor to moderate reliability, especially at low levels (PTA + 10 dB; ICC = 0.25). However, at the higher presentation levels (i.e., individual PTA + 40 dB) the standard error of the measurement was only 4% (1 word). Regarding clinical applicability, the WRS-4UFC needs to be compared to traditional speech audiometry to explore the influence of using closed-vs. open-set and forced-vs. unforced-choice test procedures on the results.

## 7 SPEECH PERCEPTION IN NOISE

The Hearing in Noise Test (HINT; Nilsson et al., 1994) is an adaptive sentence recognition test carried out with speech-shaped noise. The following assumptions are considered in HINT (based on Plomp, 1978): 1) Speech materials made of meaningful sentences yield a steep psychometric function; 2) Stationary noise with the same spectral shape as the average spectrum of the speech material makes the speech reception threshold in noise (SRT_N_) less dependent of the spectral characteristics of the speaker’s voice. Furthermore, the signal-to-noise ratio (SNR) between the target and masker is better defined across the frequency range; 3) The (SRT_N_) is independent of the absolute noise level as long as the noise level is above the “internal noise” level. Therefore, it is recommended to present the noise at least 30 dB above the “internal noise”. The internal noise is defined as the sum of the SRT in quiet of the tested listener and the SRT in noise for NH listeners, for a given speech material.

### 7.1 Methods

The Danish HINT was used as in Nielsen and Dau (2011) to obtain the SRT_N_ but in a monoaural presentation. Additionally, a 20-sentence list was presented at a fixed signal-to-noise ratio of +4 dB and scored to obtain a sentence recognition score (SScore^+4dB^). The presentation level of the noise was set between 65 and 85 dB SPL to ensure that the noise was always presented 30 dB above the individual PTA. Each ear was tested individually. All participants were tested using the same list with the same ear. Since small differences across lists were found in Nielsen and Dau (2011), this was done to ensure that all the listeners were tested with an equally difficult list. However, for the test-retest reliability study, the list and ear presented were randomized, only using lists 6-10. The listeners did not report recalling sentences from the test.

### 7.2 Results and discussion

The results of the HINT outcome measures are shown in Table 7.

**Table 7.** Summary of the results of the hearing in noise (HINT) test. The results of the speech recognition threshold in noise (SRT_N_), and sentence recognition score at +4 dB SNR (SScore^+4dB^) are shown as the mean, standard deviation, first and third quantiles (25th and 75th percentiles) for each ear.

| Outcome measure | Ear | ONH |  |  | HI |  |  |
| --- | --- | --- | --- | --- | --- | --- | --- |
|  |  | Mean (SD) | Q1 | Q3 | Mean (SD) | Q1 | Q3 |
| $SRT_N$ (dB SNR) | LE | 1.0 (0.7) | 0.4 | 1.5 | 4.1 (3.4) | 1.4 | 6.7 |
|  | RE | -0.5 (1.1) | -1.0 | 0.0 | 2.6 (3.8) | 0.0 | 4.2 |
| $SScore^{+4dB}$ (%) | LE | 85.0 (11.7) | 85 | 90 | 60.0 (26.6) | 40 | 85 |
|  | RE | 91.0 (9.6) | 90 | 95 | 62.3 (24.0) | 48.7 | 80 |

The SRT_N_ for ONH listeners were, on average, 2 dB higher than the ones reported (Nielsen and Dau, 2011). This bias was also observed in the YNH listeners. However, this might be explained by the fact that they used diotic presentation which can lead to a 1.5 dB improvement as reported by Plomp and Mimpen (1979). The results also showed a lower SRT_N_ (1.5 dB) and higher SScore^+4dB^ (4%) for the right ear in both groups of listeners. According to (Nielsen and Dau, 2011), there was a significant main effect of test list. Such differences are seen mainly for lists 1-4, which were the lists used here. Therefore, the observed interaural difference can be ascribed to a list effect, however, it might be ascribed to other factors as, for example, a right-ear advantage as the one observed in NH listeners with tinnitus (Tai and Husain, 2018). The ICC values (SRT_N_: ICC = 0.61; SScore^+4dB^: ICC = 0.57) indicated only moderate reliability of the HINT. The SRT_N_ showed an SEM = 1.02 dB, which is below the step size of the test (2 dB). The SScore^+4dB^ showed an SEM value of 7.94%, which corresponds to an error in one of the sentences. However, the reliability of the test can be improved by using an adaptive method as the one described in Wagener et al. (2003); Rønne et al. (2017) where the SRT_N_ estimation was optimized using a combination of word scoring and a maximum likelihood procedure.

## 8 SPECTRO-TEMPORAL MODULATION SENSITIVITY

A speech signal can be decomposed into spectral and temporal modulations. While speech-in-noise perception assessment leads to some confounds due to the variety of speech corpora, noise maskers, and test procedures that can all affect the results, the assessment of the contrast sensitivity of simpler sounds might be of interest for characterizing a listener’s spectro-temporal processing abilities. Bernstein et al. (2013) showed significant differences between NH and HI listeners for detecting STM in random noise. These differences corresponded to specific conditions that were also useful for the prediction of speech-in-noise performance in the same listeners. Lately, the assessment of STM sensitivity in these specific conditions gained an increasing interest due to its potential for predicting speech intelligibility (Bernstein et al., 2016; Zaar et al., 2020) and for assessing cochlear-implant candidacy (Choi et al., 2016). Here, STM sensitivity was assessed using a new test paradigm that may be more suitable for a clinical implementation. The test was performed in two conditions: a low-frequency condition (similar to the one previously used in Bernstein et al., 2016) and a high-frequency condition (Mehraei et al., 2014).

### 8.1 Methods

The stimuli were similar to those of Bernstein et al. (2016) and Mehraei et al. (2014), but a different presentation paradigm was employed. A sequence of four noises was presented in each trial. The first and third stimulus always contained unmodulated noise, whereas the second and fourth stimuli could be either modulated or unmodulated. The test was performed with a low-frequency 3-octaves wide stimulus centred at 800Hz (sSTM_8_ and fSTM_8_), and a 1-octave wide stimulus centred at 4 kHz (sSTM_4k_ and fSTM_4k_. The stimuli were presented at 75 dB sound pressure level (SPL). After the sequence was presented, the listener had to respond whether the four sounds were different (‘yes’) or the same (‘no’). Two procedures involving catch trials were evaluated. The first test (sSTM -3 dB) was a screening test consisting of 10 stimuli modulated at 20log(m) = -3 dB modulation level (ML), where m is the modulation depth, and five unmodulated ones presented in random order. The outcome measure was the listener’s contrast sensitivity (d’)^3^ in the task. The second test (fSTM) tracked the 80% threshold using a yes/no task and the single-interval adjusted matrix (SIAM; Kaernbach, 1990) paradigm. For the sSTM test, the stimulus was presented diotically whereas for the fSTM the test was presented in each ear individually in a monoaural presentation.

### 8.2 Results and discussion

The results of the spectro-temporal modulation sensitivity tests outcomes are shown in Table 8.

**Table 8.** Summary of the results of the spectro-temporal modulation sensitivity tests. The results of the screening STM sensitivity test (sSTM), and the threshold of the fast STM sensitivity test (fSTM) are shown for the low-frequency stimulus (8, because it is centered at 800 Hz) and high-frequency (4k, centered at 4kHz) as the mean, standard deviation, first and third quantiles (25th and 75th percentiles) for each ear.

| Outcome measure | Freq. Range | Ear | ONH |  |  | HI |  |  |
| --- | --- | --- | --- | --- | --- | --- | --- | --- |
|  |  |  | Mean (SD) | Q1 | Q3 | Mean (SD) | Q1 | Q3 |
| sSTM -3dB ($d'$) | LF | Bin | 2.6 (0.6) | 2.4 | 3 | 1.7 (1.3) | 0.4 | 3 |
|  | HF | Bin | 1.6 (0.8) | 1.1 | 2.4 | 0.6 (1.1) | -0.3 | 1.4 |
| fSTM (dB) | LF | LE | -7.7 (1.8) | -9 | -7.6 | -2.8 (2.1) | -3.5 | -0.8 |
|  |  | RE | -5.1 (3.1) | -7.2 | -1.6 | -1.6 (1.3) | -2 | -0.6 |
|  | HF | LE | -8.0 (2.0) | -8.6 | -6.2 | -2.6 (2.4) | -3.8 | -0.6 |
|  |  | RE | -5.6 (3.6) | -8.6 | -2.1 | -1.9 (1.5) | -2 | -1 |

The screening STM test (sSTM) shows the sensitivity in terms of d’, where the maximum value is d’ = 3, (i.e., 10 modulated and 5 unmodulated stimuli correctly detected). In the hypothetical case when all the catch trials are detected, the lowest d’ value can be -0.3. The ONH listeners showed a high sensitivity in the low-frequency condition (d’ = 2.6) and a somewhat lower sensitivity in the high-frequency condition (d’ = 1.63) corresponding to 65% correct responses. The HI listeners showed a higher variability and a lower sensitivity in the low-frequency condition (*≈* 70% correct) and substantially lower sensitivity in the high-frequency condition (0-50% correct responses). The threshold-tracking procedure (fSTM) showed results between -9 and -1.6 dB ML in the ONH group, whereas the HI listeners showed thresholds between -3.50 and -0.6 dB ML in the low-frequency condition. Although the results of the fSTM low-frequency condition were consistent with Bernstein et al. (2016), the results in the high-frequency condition showed higher thresholds than the ones in Mehraei et al. (2014). This can be ascribed to the higher presentation level used in Mehraei et al. (2014) than in the current test procedure. The fSTM showed an excellent reliability (ICC = 0.91; SEM = 0.93 dB ML) in the LF condition. However, several HI listeners were not able to complete the procedure for the HF condition. Overall, the use of the SIAM tracking procedure allowed us to obtain accurate thresholds, although additional repetitions were required, especially in the HF condition. This might be because the psychometric function for detecting the stimulus can be shallower in this condition, or because the 100% detection could not be reached even in the fully-modulated trials. Therefore, a Bayesian procedure being able to estimate the threshold and slope of the psychometric function, such as the Bayes Fisher information gain (FIG: Remus and Collins, 2008), might be more suitable for this type of test. Another reason explaining the inability of the listeners to perform the test can be ascribed to the stimulus.Zaar et al. (2018) used a longer stimulus (1s) a diotic presentation and a hearing loss compensation that ensured the audibility of the stimulus in all its frequency range. In their study, all the listeners were able to perform the tests and their sensitivity thresholds were well below the maximum value.

## 9 EXTENDED AUDIOMETRY IN NOISE (EAUD)

The extended audiometry in noise (eAUD) is a tone detection test intended to assess different aspects of auditory processing by means of a task similar to pure-tone audiometry. The tone is presented in the presence of noise and the listener has to indicate whether the tone was perceived or not. The aspects of auditory processing assessed here are 1) tone-in-noise detection, and 2) spectral and temporal resolution.

### 9.1 Tone-in-noise detection

In the pure-tone audiometry, a given patient has to detect the simple stimulus (e.g., sinusioids) in quiet aiming at estimating the hearing thresholds of the listener. A simple way to explore the supra-threshold performance is to perform a tone-in-noise detection test by presenting noise at supra-threshold levels and obtaining the masked thresholds. However, the characteristics of the noise such as bandwidth, level or inherit modulations can affect the results. Moore et al. (2000) proposed a test paradigm using a special type of noise which is able to provide the same masking in the entire frequency range, so the hearing thresholds of a NH listener would raise according to the level of the noise. This is the so-called threshold-equalizing noise (TEN). The advantage of the TEN test is that the expected masked thresholds are similar to the level of the noise (i.e., as TEN is played at 70dB per equal rectangular bandwidth (ERB), the masked threshold is expected to be at 70 dB SPL). Although this test was originally design to detect dead cochlear regions, recent evidence suggests that tone-in-noise detection can be representative of supra-threshold deficits beyond the audiogram (Schädler et al., 2020).

### 9.2 Spectro-temporal resolution

Frequency and temporal resolution are aspects of hearing that are fundamental for the analysis of perceived sounds. While NH listeners exhibit a frequency selectivity on the order of one third of an octave when using isoinput levels (from Glasberg and Moore, 1990; Eustaquio-Martín and Lopez-Poveda, 2011), HI listeners have typically broader auditory filters, leading to impaired frequency selectivity (Moore, 2007). Temporal resolution can be characterized by the ability to “listen in the dips” when the background noise is fluctuating based on the so-called masking release (Festen and Plomp, 1990). Schorn and Zwicker (1990) proposed an elaborated technique for assessing both spectral and temporal resolution using two tests: 1) Psychoacoustical tuning curves and 2) temporal resolution curves. In both cases, the task consists of detecting a pure tone that is masked by noise or another tone while the spectral or temporal characteristics of the masker are varied. Later, Larsby and Arlinger (1998) proposed a similar paradigm, the F-T test, which was successfully tested in HI listeners (van Esch and Dreschler, 2011). Here, the spectro-temporal resolution was assessed using a new test. This test is a tone-in-noise detection task consisting of three conditions as sketched in Figure 3.

**Figure 3.**
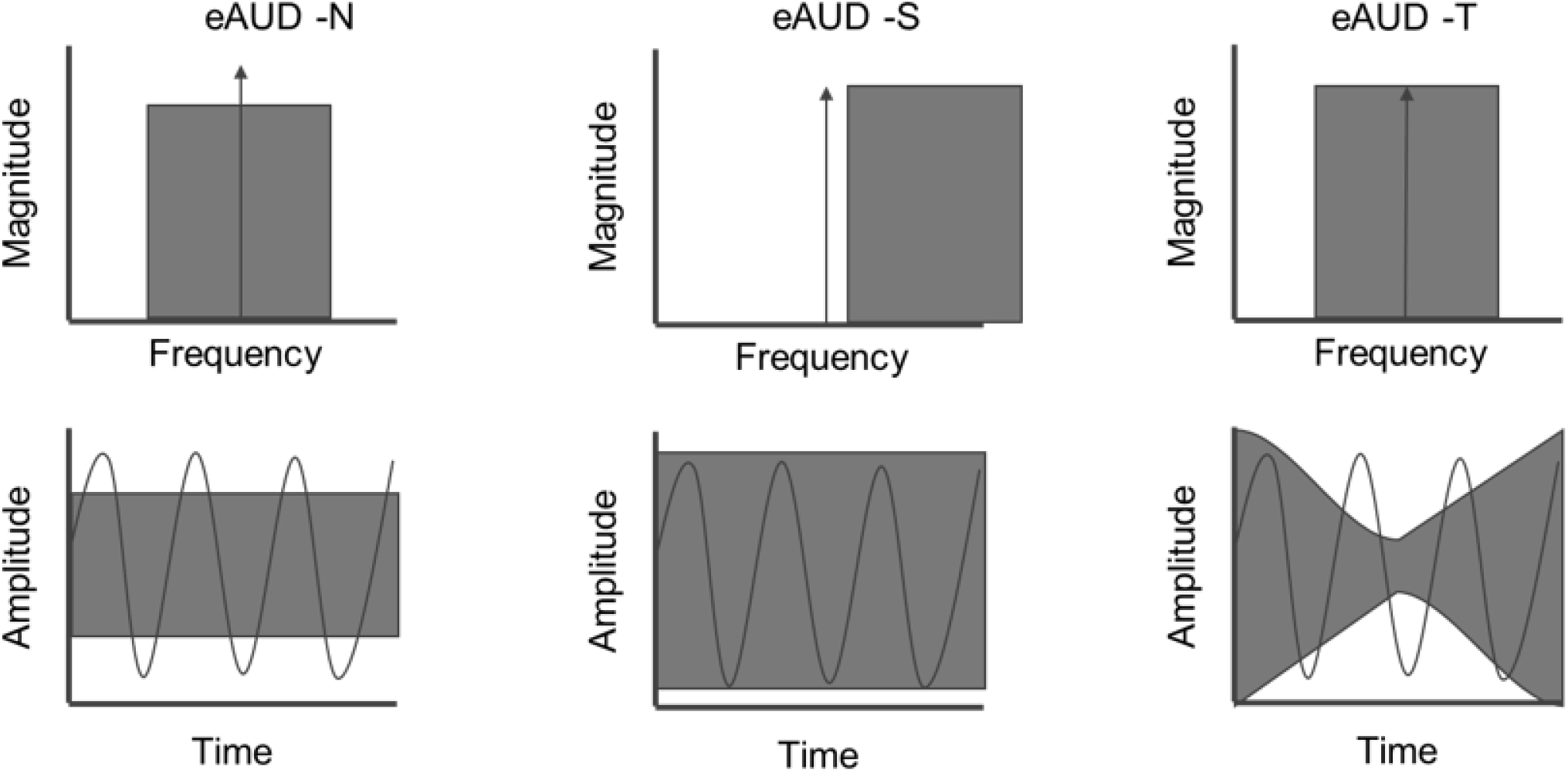
Sketch of the conditions of the spectro-temporal resolution measures of the extended audiometry in noise (eAUD). The top panel shows the spectrum of the noise and target pure-tone (delta), the bottom panel shows both signals in the time domain. Left panel: Tone in noise condition (eAUD-N). Middle panel: Spectral condition (eAUD-S). Right panel: Temporal condition (eAUD-T).

1. eAUD-N: The tone is embedded in a 1-octave-wide threshold equalizing noise (TEN-HL; Moore, 2001). Because of the properties of the TEN-HL, the tone detection threshold is comparable to the level of the noise in dB HL.
2. eAUD-S: The tone is embedded in a TEN that has been shifted up in frequency. In the spectral domain, this yields spectral unmasking of the tone, so the detection threshold is lower than in eAUD-N.
3. eAUD-T: The tone is embedded in a temporally-modulated noise with the same spectral properties as the one in eAUD-N. In the temporal domain, the modulations of the noise yield temporal unmasking, so the tone can be detected in the dips.

The outcome measures were focused on the temporal and spectral benefits expected in the eAUD-S and eAUD-T conditions compared to the eAUD-N condition. While in the noise condition (eAUD-N) the threshold is expected to be approximately at the level of the noise, in the temporal and spectral conditions the thresholds should be obtained at a lower level, showing temporal masking release (TMR) and spectral masking release (SMR)

### 9.3 Methods

The procedure used here was a yes/no task using a SIAM procedure (Kaernbach, 1990) similar to the one used in the eAUD-HF. Here, a TEN was presented together with a warble tone. If the target was detected, the target-presentation level is decreased according to a given step size; if it was not detected, the level is increased. If the stimulus was not presented (catch trial) but the listener provided a positive response, the level is decreased compared to the previous trial. As in the eAUD-HF, for each run, the threshold of each trial was calculated as the average of the last four reversals. The noise was presented at 70 dB HL. The low-frequency condition (LF) corresponds to the detection of a 0.5-kHz warble tone, whereas the high-frequency (HF) condition corresponded to a 2-kHz warble tone. The final threshold was calculated as the mean threshold of two repetitions. In the eAUD-S condition, the center frequency of the noise was *f*_*c*,noise_ = 1.1*f*_tone_. In the eAUD-T condition, the modulation frequency of the noise was set to, *f*_*m*_ = 4 Hz. The outcome measures of the eAUD are 1) the tone-in-noise threshold (TiN), 2) the temporal masking release (TMR), and 3) the spectral masking release (SMR).

### 9.4 Results and discussion

The results of the extended audiometry in noise outcomes are shown in Table 9.

**Table 9.** Summary of the results of the extended audiometry in noise (eAUD). The results of the tone-in-noise (TiN), temporal masking release (TMR) and spectral masking release (SMR) are shown for the low-frequency condition (LF; 500Hz) and high-frequency condition (HF; 2kHz) as the mean, standard deviation, first and third quantiles (25th and 75th percentiles) for each ear.

| Outcome measure | Freq. Range | Ear | ONH |  |  | HI |  |  |
| --- | --- | --- | --- | --- | --- | --- | --- | --- |
|  |  |  | Mean (SD) | Q1 | Q3 | Mean (SD) | Q1 | Q3 |
| TiN (dB HL)<br>eAUD-N | LF | LE | 70.4 (4.5) | 68 | 71.5 | 71.8 (2.6) | 70.2 | 73.2 |
|  |  | RE | 69.2 (4.6) | 65.2 | 72.5 | 72.0 (2.8) | 69.6 | 74.3 |
|  | HF | LE | 71.1 (2.5) | 69.7 | 72.7 | 74.7 (3.4) | 72.5 | 76.1 |
|  |  | RE | 70.8 (3.6) | 70.5 | 71.7 | 74.2 (3.1) | 72 | 76.2 |
| TMR (dB)<br>eAUD (N-T) | LF | LE | 7.5 (3.4) | 6 | 7.5 | 7.7 (4.0) | 6.1 | 10.1 |
|  |  | RE | 5.2 (3.3) | 4 | 7.6 | 8.3 (2.7) | 6.5 | 10.3 |
|  | HF | LE | 13.0 (0.6) | 12.7 | 13.2 | 7.9 (5.0) | 5 | 11.6 |
|  |  | RE | 10.7 (3.1) | 9.1 | 10.2 | 8.1 (5.2) | 5.1 | 10.7 |
| SMR (dB)<br>eAUD (N-S) | LF | LE | 19.3 (3.6) | 16.5 | 21.7 | 19.6 (17.7) | 17.7 | 23.2 |
|  |  | RE | 18.8 (4.6) | 17 | 21.2 | 20.0 (5.2) | 16.5 | 23.8 |
|  | HF | LE | 26.8 (4.5) | 27.5 | 29 | 19.3 (9.5) | 12.1 | 26.3 |
|  |  | RE | 27.2 (3.7) | 26.2 | 29.5 | 19.5 (9.9) | 12 | 26.8 |

The TiN showed a larger variance for the ONH group (SD = 4.5 dB HL) at low frequencies. The detection thresholds were in line with previous work with thresholds close to the noise presentation level (70 dB HL) (Vinay et al., 2017). The TMR shown by the NH group was larger at high frequencies (10 dB) than at low frequencies (7 dB). The HI group showed, on average, similar TMR only at low frequencies. The SMR shown by the ONH listeners was 19 dB for low frequencies and 26 dB for high frequencies. In contrast, for the HI listeners, the SMR was 7 dB lower only in the high-frequency condition. The reliability of the eAUD was moderate for most of the conditions (ICC*≤* 0.75). The eAUD-S at low frequencies showed good reliability (ICC = 0.85; SEM = 1.78 dB). The masking release estimates showed good reliability only for the high-frequency condition. The reason for this might be that masking release is a differential measure, and the cumulative error is, therefore, higher than that of each individual measure. The reduced reliability can be explained to some extent by the method used. To have a similar procedure as in pure-tone audiometry, the parameters of the SIAM tracking procedure were set accordingly. However, this made the test challenging and the listeners consistently missed several catch trials. Thus, extra trials were required to improve measurement accuracy, especially in the eAUD-N condition. Furthermore, the standard error of the measurement was in most cases larger than the final step size (2 dB). As in the case of the fSTM, a different procedure, such as Bayesian adaptive methods, might increase measurement reliability.

## 10 BINAURAL PROCESSING ABILITIES

Binaural hearing is useful for sound localization and the segregation of complex sounds (Darwin, 1997). Interaural differences in level or timing are processed for spatial hearing purposes in the auditory system. In the case of hearing loss, the neural signal at the output of the cochlea can be degraded which may lead to reduced binaural abilities typically connected to temporal fine structure (TFS) processing. Based on a method of estimating the upper-frequency limit for detecting an interaural phase difference (IPD) of 180° (IPD_fmax_ Ross et al., 2007; Neher et al., 2011; Santurette and Dau, 2012), Fü llgrabe and Moore (2017) recently proposed a refined test as a feasible way to evaluate TFS sensitivity. This paradigm was used in recent research that suggested that IPD_fmax_ might be related to non-auditory factors (Strelcyk et al., 2019) and affected by factors beyond hearing loss, such as musical training (Bianchi et al., 2019). Therefore, the IPD_fmax_ might be a task that requires auditory and non-auditory processing abilities beyond TFS sensitivity.

In contrast, binaural pitch detection assesses binaural processing abilities in a different manner. This test requires the detection of pitch contours embedded in noise, which are diotically or dichotically evoked. While the diotic condition can be resolved monoaurally, the dichotic condition requires the binaural processing abilities to be sufficiently intact to detect the contour. Previous studies showed that some listeners were unable to detect binaural pitch, regardless of the audiometric configuration (Sanchez-Lopez et al., 2018; Santurette and Dau, 2012). Therefore, it was of interest to compare the results of these two binaural processing tests.

Another approach for evaluating the binaural processing abilities is assessing binaural masking release (Durlach, 1963), which has been used in several studies (e.g. Neher, 2017; Strelcyk and Dau, 2009) and implemented in some commercial audiometers (Brown and Musiek, 2013). In this paradigm, a tone-in-noise stimulus is presented in two conditions: (1) a diotic condition where the tone is in phase in the two ears, and (2) a dichotic condition where the tone is in antiphase in the two ears. The difference between the two yields the benefit for tone detection due to binaural processing, the so-called binaural masking release (BMR).

### 10.1 Methods

The maximum frequency for detecting an IPD of 180° with pure-tones was obtained using a 2-AFC tracking procedure similar to the one used in Fü llgrabe and Moore (2017). The stimuli were presented bilaterally in both ears as two sequencies of four tones. One sequence contained an ABAB sequence, where A means a dichotic presentation and B an IPD of 180° between the tones presented to each ear, and the other an AAAA sequence. A positive response (detection) increased the frequency of the tone, and a negative response a decrease of the frequency. Although the stimuli duration and procedure were similar, the step size used here was slightly different, starting with steps of 2/3 octave and decreasing to a final step size of 1/6 octave in each reversal. The last six reversals were used for estimating the threshold. The frequency threshold (IPD_fmax_) was obtained from the average of two runs. Binaural pitch detection scores were obtained using a clinical implementation of the test proposed by Santurette and Dau (2012). A 3-minute sequence of noise was presented bilaterally. Ten diotic and ten dichotic pitch contours, embedded in the noise, had to be detected by the listener. The tones forming the pitch contours were generated by adding frequency-specific IPDs to the presented noise (Cramer and Huggins, 1958). The outcome measure of the binaural pitch test was the percentage score of detecting the dichotic pitch contours only, averaged across two repetitions (BP20). The BMR was assessed using the same method as the extended audiometry. Two measurements were required: 1) tone-in-noise detection presented diotically (S_0_N_0_) and tone-in-noise detection presented dichotically, i.e., with the tone in anti-phase across the two ears (S_*π*_N_0_).

### 10.2 Results and discussion

The results of the tests assessing binaural processing abilities are shown in Table 10.

**Table 10.** Summary of the results of the binaural processing abilities tests. The results of the maximum frequency for IPD detection (IPD_fmax_), Binaural Pitch detection scores (BP_20_) and the binaural masking release (BMR) are shown as the mean, standard deviation, first and third quantiles (25th and 75th percentiles) for each ear.

| Outcome measure | Ear | ONH |  |  | HI |  |  |
| --- | --- | --- | --- | --- | --- | --- | --- |
|  |  | Mean (SD) | Q1 | Q3 | Mean (SD) | Q1 | Q3 |
| $IPD_{fmax}$ (kHz) | Bin | 0.76 (0.26) | 0.59 | 0.98 | 0.69 (0.27) | 0.52 | 0.88 |
| Bin Pitch 20 (%) | Bin | 87.5 (25.0) | 87.5 | 100 | 80.7 (30.9) | 70 | 100 |
| BMR (dB)<br>($S_0N_0 - S_\pi N_0$) | Bin | 16.5 (4.7) | 13.5 | 17.5 | 14.7 (4.6) | 12.2 | 17.5 |

The listeners in the ONH and HI groups showed IPD_fmax_ thresholds around 700 Hz with a standard deviation (*≈* 270 Hz) and interquartile range (*≈* 370 Hz) similarly in both groups. These results are in line with the ones reported in Füllgrabe and Moore (2017). The IPD_fmax_ test showed excellent reliability (ICC = 0.95; SEM = 65.4 Hz), and the median time needed for two repetitions was 10 minutes. This suggests that IPD_fmax_ is a reliable measure of binaural processing abilities that can reveal substantial variability among both NH and HI listeners, which is valuable for highlighting individual differences among patients. The overall results from the binaural pitch test for the NH listeners showed *>* 87.5% correct detection, whereas the HI listeners’ results showed a higher variability with an interquartile range from 70-100%. The test showed excellent reliability (ICC = 0.98; SEM = 4%). Listeners reported a positive experience due to the test being short and easy to understand. The BMR shown by both groups was around 15 dB, as expected from previous studies (Durlach, 1963).

## 11 EXPLORATORY ANALYSIS

The collection of tests included in the test battery was intended to explore different and potentially independent aspects of hearing to obtain an auditory profile with controlled interrelations among the tests. A factor analysis performed in the HEARCOM study (Vlaming et al., 2011) based on data from 72 HI subjects revealed auditory dimensions: 1) high-frequency processing, 2) audibility, 3) low-frequency processing and 4) recruitment. In the current study, the results of the behavioural tests were analysed further in order to explore possible interrelations between the various outcome measures.

### 11.1 Methods

First, the data were pre-processed as in Sanchez-Lopez et al. (2018) to reduce the number of variables. The outcome variables of the frequency-specific tests were divided into LF (*≤* 1 kHz) and HF (¿1 kHz) variables. This decision was supported by a correlation analysis performed on the complete set of outcome variables, where the outcomes corresponding to 2, 4 and 6 kHz as well as the ones corresponding to 0.25, 0.5 and 1 kHz were highly intercorrelated. For the tests performed monaurally, the mean of the two ears was taken as the resulting outcome variable. The resulting dataset (BEAR3 dataset^1^) contained 26 variables, divided into six groups corresponding to the six aspects of auditory processing considered here. The exploratory analysis consisted of a correlation analysis using Spearman correlations and factor analysis. The factor analysis was performed using an orthogonal rotation (“varimax”) and the method of maximum likelihood. The number of components was chosen to use parallel analysis, the resulting number of components was four.

### 11.2 Results

Figure 4 shows the results from the correlation analysis performed on the BEAR3 dataset. For convenience, the absolute value of the correlation was used when visualizing the data to show the strength of the correlation. The circles on the left-hand side of the figure depict significant correlations (*p <* 0.00001), and the correlation values are presented on the left-hand side of the figure. Two groups of correlated variables can be observed. The upper-left corner shows variables related to LF processing (dynamic range, the slope of the loudness function, and hearing thresholds) and speech intelligibility in quiet. The bottom-right corner shows a larger group of correlated variables including HF processing, speech intelligibility in noise, and spectro-temporal resolution at high frequencies. The variables that are not significantly interrelated are shown in the middle part of Figure 3, including the three variables related to binaural processing abilities (IPD_fmax_, BP20 and BMR) which were not significantly correlated to each other. The speech reception threshold in quiet (SRT_Q_) and the STM detection were correlated to various variables such as tone-in-noise detection, HF spectro-temporal resolution, LF hearing thresholds and speech-in-noise perception.

**Figure 4.**
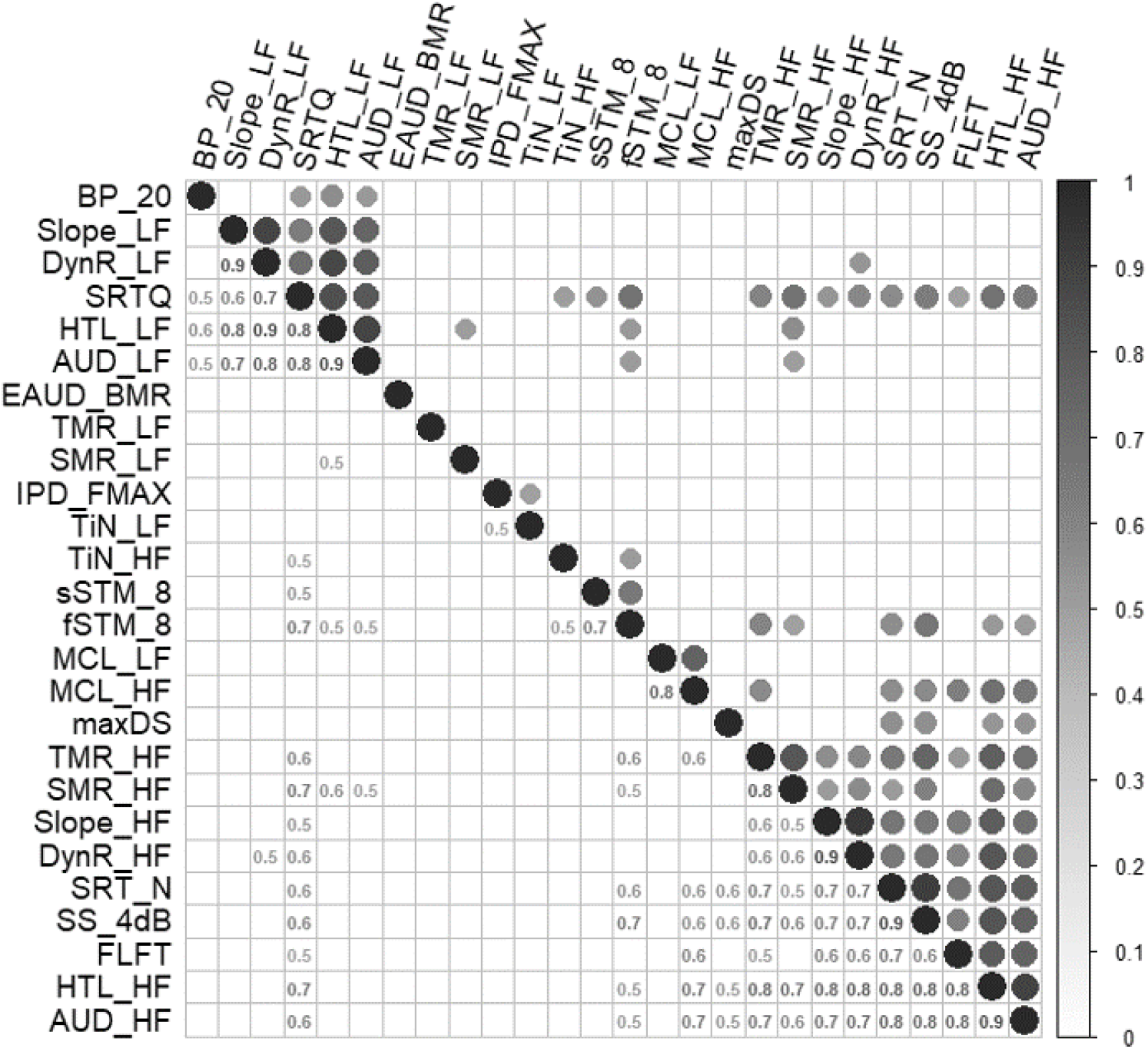
Correlation plot of the data set BEAR3. The upper part shows the significantly correlated variables as coloured circles. The lower panel shows the numeric correlation value.

The four factors resulting from the factor analysis showed 63% of explained cumulative variance. The variables with higher loadings (*>* 0.65) for each of the factors are shown in Table 11. The first factor, in terms of the amount of variance explained (19%), was associated with LF loudness perception and speech intelligibility in quiet, whereas the second factor (18% of variance explained) was associated with HF loudness perception. Despite loudness perception being associated with the first and second factor, the MCL was associated, both at high and low frequencies, with the third factor, while the fourth factor was associated with speech intelligibility in noise.

**Table 11.** Variables correlated to the four latent orthogonal factors resulting from the factor analysis with the method of maximum likelihood (ML). Columns are sorted in terms of the variance explained by each factor.

|  | *ML2(19%)* | *ML1(18%)* | *ML3(14%)* | *ML4(12%)* |
| --- | --- | --- | --- | --- |
| *HTL_LF* | 0.93 |  |  |  |
| *DynR_LF* | -0.9 |  |  |  |
| *AUD_LF* | 0.82 |  |  |  |
| *Slope_LF* | 0.81 |  |  |  |
| *SRTQ* | 0.67 |  |  |  |
| *DynR_HF* |  | -0.93 |  |  |
| *Slope_HF* |  | 0.82 |  |  |
| *HTL_HF* |  | 0.79 |  |  |
| *AUD_HF* |  | 0.73 |  |  |
| *MCL_HF* |  |  | 0.92 |  |
| *MCL_LF* |  |  | 0.85 |  |
| *SRT_N* |  |  |  | 0.77 |
| *SScore_4dB* |  |  |  | -0.78 |

## 12 GENERAL DISCUSSION

The first goal of the present study was to collect data of a heterogeneous population of HI listeners, reflecting their hearing abilities in different aspects of auditory processing. The current study was motivated by the need for a new dataset to refine the data-driven approach for auditory profiling. The dataset should contain a representative population of listeners and outcome measures (Sanchez-Lopez et al., 2018) to allow a refined definition of the two types of auditory distortions and to identify subgroups of listeners with clinical relevance. To refine the data-driven auditory profiling, the BEAR3 dataset fulfils all the requirements discussed in Sanchez-Lopez et al. (2018). Other datasets containing a large number of listeners (e.g., Gieseler et al., 2017; Rönnberg et al., 2016) or physiological measures (e.g., Kamerer et al., 2019) could also be interesting for complementing the auditory profiling beyond auditory perceptual measures.

### 12.1 Relationships across different aspects of auditory processing

The proposed test battery considers outcomes divided into six dimensions of auditory processing. One of the objectives of the study was to investigate the interrelations of different dimensions and measures. The present analysis showed two interesting findings. First, the correlation analysis shows two clusters of variables related to either low- or high-frequency audiometric thresholds. Speech-in-noise perception was associated with high-frequency sensitivity loss, temporal, and spectral masking release, whereas speech-in-quiet was correlated with both low- and high-frequency hearing loss. Several outcomes were not interrelated, especially the outcomes associated with binaural processing abilities. Second, factor analysis yielded latent factors related to low- and high-frequency processing, most comfortable level and speech in noise. Vlaming et al. (2011) showed four dimensions in the factor analysis of the HEARCOM project data corresponding to high-and low-frequency spectro-temporal processing, MCL and recruitment. In contrast, the current study showed that the slopes of the loudness growth, both at low and high frequencies, were not interrelated and contributed to the first and second latent factors. Additionally, the speech-in-noise test performed in HEARCOM was associated with the low-frequency processing, whereas, in the present study, speech-in-noise dominates the fourth factor and is significantly correlated with high frequencies. The reason for this discrepancy might be the use of different types of noise and test procedures in the two studies.

Overall, the data of the present study seem to be dominated by the audiometric profiles, with low- and high-frequency processing reflecting the main sources of variability in the data. However, binaural processing abilities, loudness perception and speech-in-noise outcomes showed a greater contribution to the variability of the supra-threshold measures than spectro-temporal processing outcomes.

### 12.2 Effects of the participant’s cognitive abilities

In this study, only auditory tests were considered. Indeed, some of the tests were quite demanding, which might have affected the results of listeners with reduced cognitive abilities. Here, only the age could be indicative of a likely cognitive decline, but it can not confirm or deny this. In terms of age, we observed a significant effect on the results of the IPD and tone-in-noise tests. However, a thorough analysis on the effect of age was not carried out with the existing data. A more interesting approach would be to replicate this study including cognitive tests assessing executive functions, working memory or attention span with the aim of including a heterogeneous group of listeners with various cognitive abilities. Such a study could potentially assess both hearing and cognitive abilities towards a feasible test battery that can be included in the audiological assessment, either in or out of the clinic.

### 12.3 Towards clinical feasibility of the tests

The test-retest reliability of the test battery was investigated based on the results of a subset of listeners who participated 2-5 months after the first visit. The analysis was based on the ICC and the SEM. Some of the tests, such as IPD_fmax_, binaural pitch and eAUD-HF (FLFT) showed good to excellent test-retest reliability with all ICC values above 0.9, while other tests, such as the extended audiometry in noise and speech intelligibility in quiet, showed poor reliability. The selected tests were conducted in two sessions and the total time was, on average, three hours including the instructions and interview. In realistic clinical setups, a subset of tests with high reliability and a reasonably low difficulty would need to be prioritized. For a clinical version of the test battery, other tracking procedures such as Bayesian Functional information (Remus and Collins, 2008) might be adopted to improve the reliability and time-efficiency in some tasks such as STM and tone detection in noise. Moreover, if time-efficiency is crucial, testing some aspects of auditory processing out of the clinic, as other proposed test batteries for auditory research (Lelo De Larrea-Mancera et al., 2020), might be a solution for completing the patient’s hearing profile. The use of speech-in-noise tests can be a useful tool for the characterization of the listener’s hearing deficits that can be performed under different conditions, including monaural, binaural, unaided and aided stimuli presentations. While here the tests were performed monaurally and unaided, a binaural condition as well as at least one aided measure (i.e., with hearing aids), could also be included in clinical practice. A clinical test battery with the subset of tests that showed a good or excellent test-retest reliability should be evaluated in a large scale study. In this study, we explored the use of an extended audiometry using the same test procedure for assessing high-frequency audibility (eAUD-HF), tone-in-noise detection (eAUD-N), spectro-temporal resolution (eAUD-S and eAUD-T), and binaural processing abilities. This procedure can be further explored and be performed by a hearing-care professional rather than in the current experimental setup. However, if the goal is to accurately estimate the hearing deficits of the patient, the test battery should include several aspects of auditory processing and provide detailed information on the supra-threshold deficits of the patient. The tests that showed potential for the clinical implementation were ACALOS, HINT, fSTM (only the LF condition), Binaural Pitch and IPD_fmax_. Such a test battery could serve to identify a clinically relevant subset of patients (auditory profiles) that may benefit from specific types of hearing rehabilitation towards a “stratified approach” (Lonergan et al., 2017) for audiology practice.

## 13 CONCLUSION

The current study has shown the rationale behind the BEAR test battery and the selected tests for characterizing hearing deficits in listeners with various hearing abilities. The analysis of the data showed that a reduced BEAR test battery has the potential for clinical implementation, providing relevant and reliable information reflecting several auditory domains. The proposed test battery showed good reliability, was reasonably time-efficient and easy to perform. The implementation of a clinical version of the test battery is to be evaluated in future research, e.g., in a larger field study to further refine the auditory profiling approach. Moreover, the current data has been already analyzed for the purpose of auditory profiling (Sanchez Lopez et al., 2020), showing the potential of this test battery for hearing rehabilitation.

## Data Availability

The data that support the findings of this study are openly available in Zenodo at http://doi.org/10.5281/zenodo.3459579.

## 14 ETHICS STATEMENT

All the patients received written informed consent. The study was approved by the Science-Ethics Committee for the Capital Region of Denmark H-16036391.

## AUTHOR CONTRIBUTIONS

Author contributions according to [CRediT](http://credit.niso.org/) (Contributor Roles Taxonomy). RSL: conceptualization, methodology, software, validation, formal analysis, investigation, data curation, visualization and writing-original draft and editing. SGN: investigation, validation, resources, formal analysis, visualization, writing-Original draft. ME: methodology, investigation, resources, writing-review. MF and FB: conceptualization, methodology, supervision and writing-editing. MW and OC: investigation, resources and writing-review. TN: methodology, supervision, project administration and writing-review. SS and TD: conceptualization, methodology, supervision, project administration and funding acquisition. All authors contributed to the article and approved the submitted version.

## ACKNOWLEDGMENTS

We thank the staff from OUH, BBH and HEA, especially JH Schmidt, SS Houmøller, E Kjærbøl, RS Sørensen, and the student helpers from the MSc of Audiology at SDU. We also want to show our gratitude to all the participants in the study. We want to thank I. Holube and two annonymous reviewers for their comments in an earlier version of this manuscript, as well as SD Ewert for the AFC framework. This work was supported by Innovation Fund Denmark Grand Solutions 5164-00011B (Better hEAring Rehabilitation project), Oticon, GN Resound, Widex and other partners (Aalborg University, University of Southern Denmark, the Technical University of Denmark, Force technology, Aalborg, Odense and Copenhagen University Hospitals).The funding and collaboration of all BEAR partners are sincerely acknowledged.

## DATA AVAILABILITY STATEMENT

The datasets generated from this study can be found in the Zenodo repository: https://www.doi.org/10.5281/zenodo.3459579 A working version of the Test Battery can be shared upon a reasonable request to the corresponding author. This study is part of the PhD thesis entitled ”Clinical auditory profiling and profile-based hearing-aid fitting” and available as the volume 43 of the Contributions to Hearing Research at the Technical University of Denmark.

## Footnotes

1 The BEAR3 available at Zenodo contains an observation labelled ’0’, which corresponds to the results of one of the examiners and it is not used in the present analysis

2 The factors identified correspond to the authors’ conclusions based on cited references. For example, Johannesen et al. (2016) identified the basilar membrane compession as a predictor of speech intelligibility in stationary noise and temporal processing as a predictor of speech-in-speech intelligibility. Desloge et al. (2017); Oxenham and Simonson (2009); Rhebergen et al. (2006) identified the audibility of the soft speech sounds in the presence of fluctuating maskers as a crucial factor for speech intelligibility.

3 Despite other listeners presented PTA *≤* 22 dB, the individual thresholds did not fulfil this criteria.

3 d’ was defined as 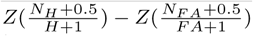 where Z refers to the z-score transformation, H is the total number of target presentations and FA the total number of catch trials.

